# Hepatitis B virus resistance to tenofovir: fact or fiction? A synthesis of the evidence to date

**DOI:** 10.1101/19009563

**Authors:** Jolynne Mokaya, Anna L McNaughton, Phillip A Bester, Dominique Goedhals, Eleanor Barnes, Brian D Marsden, Philippa C Matthews

## Abstract

**Background:** Tenofovir (TFV) is a widely used antiviral treatment for chronic hepatitis B virus (HBV) infection. There is a high genetic barrier to the selection of TFV resistance-associated mutations (RAMs), but the distribution and clinical significance of TFV RAMs are not well understood, and the topic remains contentious. We here present assimilated evidence for putative TFV RAMs with the aims of cataloguing and characterising mutations that have been reported, and starting to develop insights into the mechanisms of resistance and potential clinical significance.

**Methods:** We carried out a systematic literature search in PubMed to identify clinical, *in vitro* and *in silico* evidence of TFV resistance. The structure of HBV reverse transcriptase (RT) has not been solved; we therefore compared HBV RT to the crystal structure for HIV RT to map the likely sites of RAMs.

**Results:** We identified a ‘long-list’ of 37 putative TFV RAMs in HBV RT, occurring within and outside sites of enzyme activity, some of which can be mapped onto a homologous HIV RT structure. Based on quality and quantity of supporting data, we generated a ‘short-list’ of nine sites that are supported by the most robust evidence. Most resistance arises as a result of suites of multiple RAMs. Other factors including adherence, viral load, HBeAg status, HIV coinfection and NA dosage may also influence viraemic suppression.

**Conclusion:** There is emerging evidence for polymorphisms that may reduce susceptibility to TVF. A better understanding of HBV drug resistance is imperative to optimise approaches to public health elimination targets.

## INTRODUCTION

Nucleot(s)ide analogues (NA) are the most widely used antiviral treatments for chronic HBV (CHB) infection (1), with tenofovir (TFV) being safe, cheap, and widely available agent. NA agents inhibit the action of HBV reverse transcriptase (RT), acting as DNA chain terminators. NA therapy can be effective in suppressing HBV viraemia, thus reducing the risks of inflammation, fibrosis and hepatocellular carcinoma (HCC) as well as lowering the risk of transmission (1). However, NAs are not curative due to the persistent intracellular hepatic reservoir of HBV covalently closed circular DNA (cccDNA). Long-term administration is therefore typically required (2), with a potential risk of selection of resistance associated mutations (RAMs) in the virus (3,4). RAMs are mostly likely to arise in the context of high viral replication, arising as a result of the error prone RT enzyme (1).

Lamivudine (3TC), telbivudine (LdT) and adefovir (ADV) have been phased out of use in HBV management, mainly due to the predictable selection of RAMs over time (1). The best recognised 3TC RAM arises at RT-M204, representing the second position of the tyrosine-methionine-aspartate-aspartate (‘YMDD’) motif in viral RT (1,4). Resistance to TFV, formulated either as tenofovir disoproxil fumarate (TDF) or tenofovir alafenamide (TAF) (**Suppl Fig 1**), remains controversial. Unlike other NAs, TFV has a high genetic barrier to resistance (1), corroborated by studies that report no resistance after many years of treatment (5). An on-line tool, ‘geno2pheno hbv’, lists only one position (RT N236T) in association with reduced TFV susceptibility, while other reports also include A181T/V (6). However, there are emerging reports of a wider range of amino acid substitutions that are associated with reduced TFV sensitivity, described in both treatment-experienced and treatment-naïve individuals with CHB (7,8).

**Fig 1:**
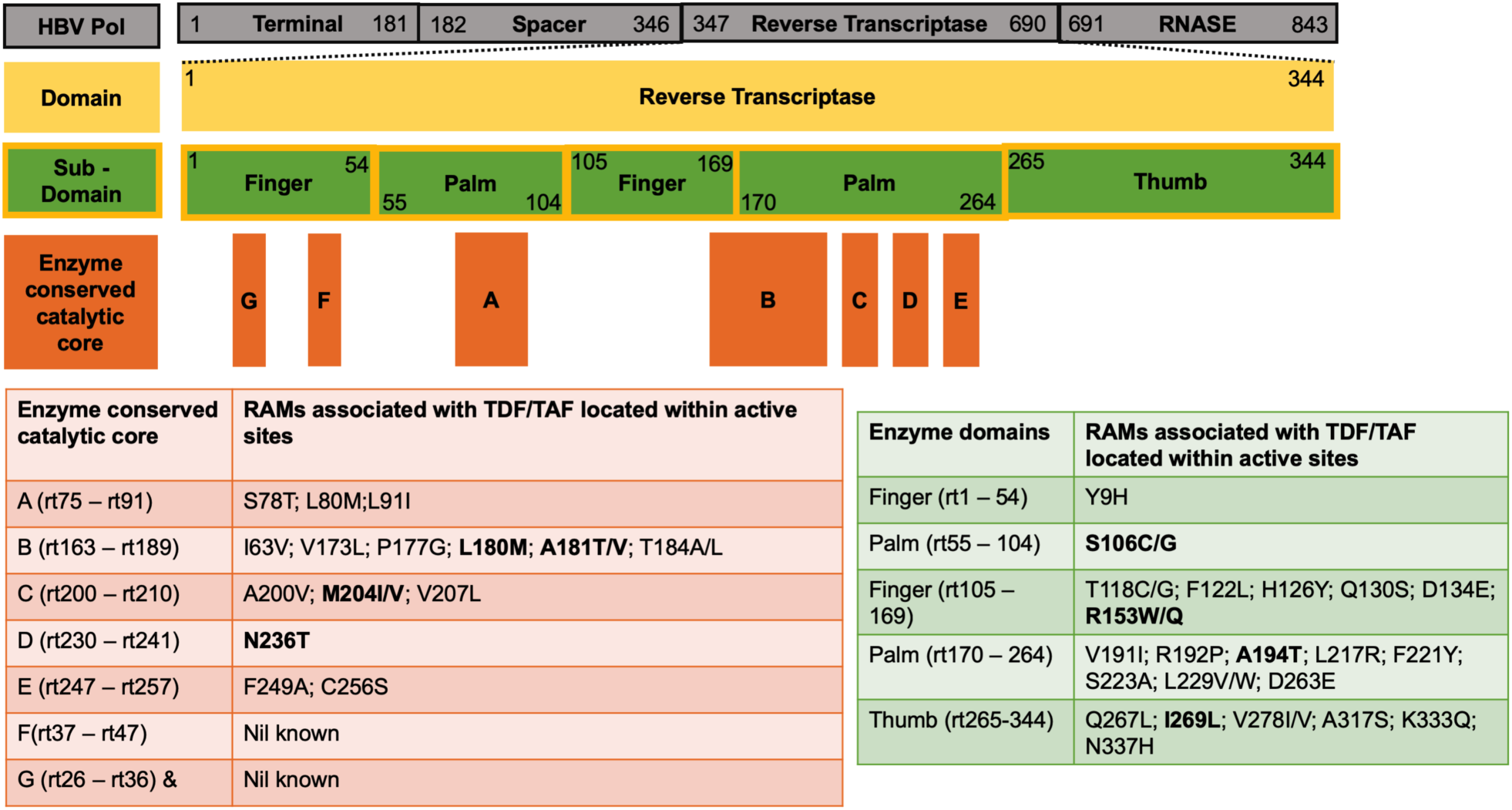
Mutations associated with tenofovir (TFV) resistance located within and outside the active sites of the HBV RT enzyme. Polymerase numbering shown in the grey bars is based on genotype A sequence (accession number X02763). Yellow bar represents RT; green bars represent subdomains which are designated finger, palm and thumb; orange rectangles represent active sites of the enzyme referred to as regions A-G. Mutations associated with TFV resistance (n=37 sites) are listed according to their location within active sites of the enzyme (orange table) or outside active sites (green table). The sites shown in bold represent the nine mutations in our short-list with best literature support (evidence summarized in Table 1). Note that in most cases, individual mutations are unlikely to be sufficient to mediate resistance, and a resistant phenotype arises only as a result of combinations of ≥2 polymorphisms.

There is some degree of homology between the sequence, structure and function of HIV and HBV RT enzymes, explaining why certain NAs are active against both viruses (9). Although no crystal structure has been resolved for HBV RT, some studies have modelled this enzyme based on the HIV crystal structure (9–11), suggesting that insights into HBV drug resistance mechanisms might be inferred from what is known about HIV.

**Table 1:**
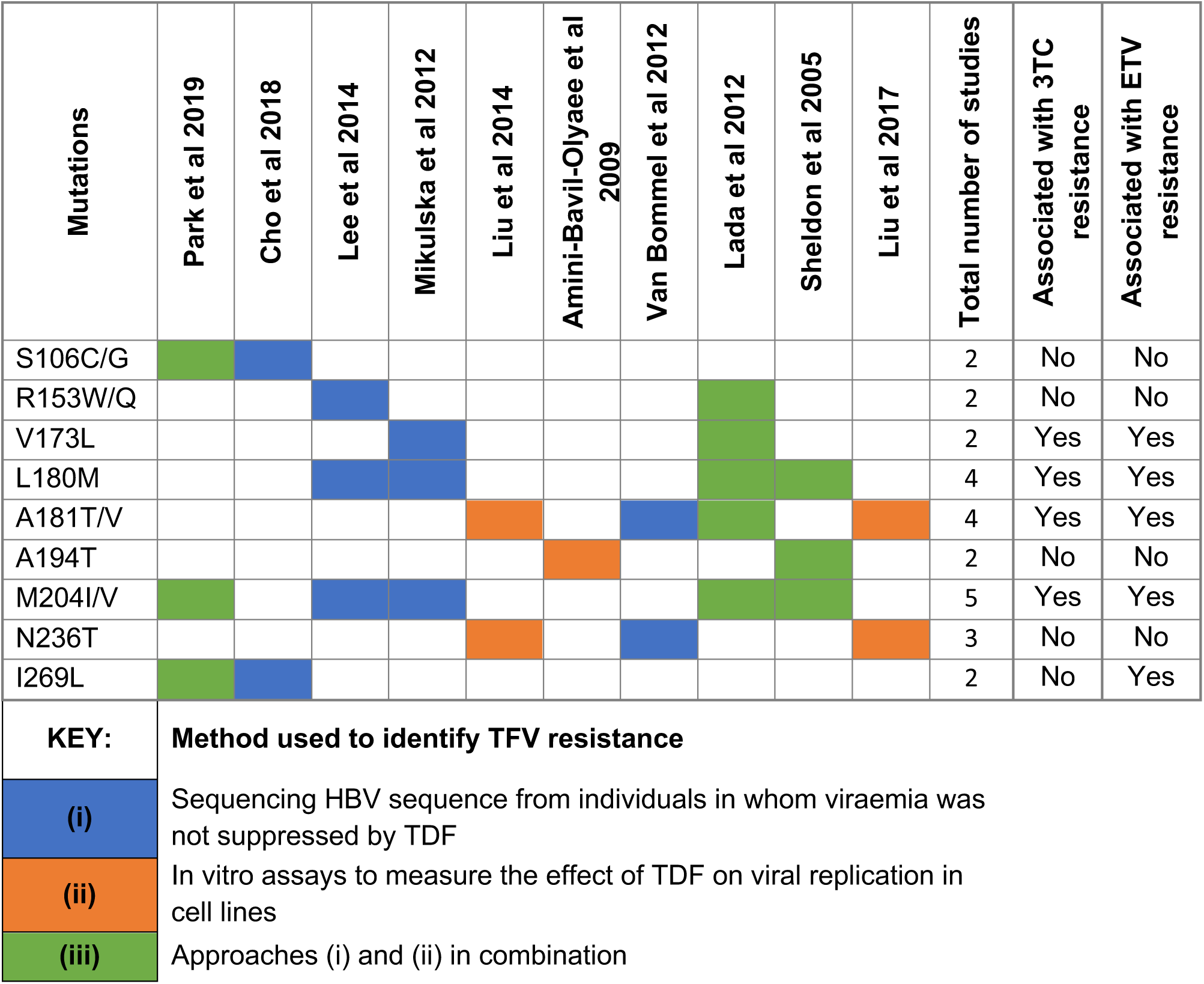
Mutations most commonly reported in association with tenofovir resistance in HBV, identified from a systematic literature review. This table reports a ‘short-list’ of mutations identified in two or more different studies (7,8,20,24–26,28–30,32). The color indicates the method(s) used to determine drug resistance. All positions are in HBV RT, listed in numerical order. The ‘long-list’ of 37 mutations identified in 15 included studies is reported in Suppl Table 3.

A better understanding of the role of NA therapy in driving HBV elimination at a population level is crucial to underpin efforts to move towards international targets for elimination by the year 2030 (3,12). For populations in which HIV and HBV are both endemic, as exemplified by many settings in sub-Saharan Africa, there are particular concerns about drug resistance in HBV, given the widespread population exposure to TDF as a component of first-line antiretroviral therapy (ART) for HIV (3). In order to progress towards these targets, many more people will need to be treated in the decade ahead.

We have therefore undertaken a systematic approach to assimilate the current evidence for the development of clinical or virological HBV breakthrough during TFV therapy. The evidence on this topic is not currently sufficiently advanced to underpin definitive conclusions regarding specific genetic signatures that underpin TVF resistance, or the extent to which these are significant in clinical practice. However, we add to the field by providing a comprehensive summary of relevant publications, together with a quality appraisal of the evidence. We used this process to assimilata a ‘long-list’ (all putative TFV RAMs) and a ‘short-list’ (a refined catalogue containing only the polymorphisms most robustly supported by existing data). We highlight gaps in the existing data and the urgent need for more research.

## METHODS

### Search strategy and quality appraisal

We undertook a systematic search of PubMed and Scopus in February 2019, using PRISMA criteria (**Suppl Fig 2**). We used the search terms (“*Hepatitis B virus” [Mesh] OR “hepatitis b” OR HBV*) AND (*Tenofovir OR TDF* OR *TAF* OR *“Tenofovir alafenamide”* OR “*Tenofovir Disoproxil Fumarate*”) *AND* (*resista* OR drug muta* OR DRMs OR RAMs OR viremia OR replica\**). We reviewed the titles and abstracts matching the search terms and included those reporting virological HBV breakthrough after exposure to TFV, only including studies that presented original data and had undergone peer review. We used the Joanna Briggs Institute Critical Appraisal tool checklist to assess for quality of case reports (13). For assessment of quantitative studies, we used the BMJ adapted Quality Assessment Tool for Quantitative Studies (14).

**Fig 2:**
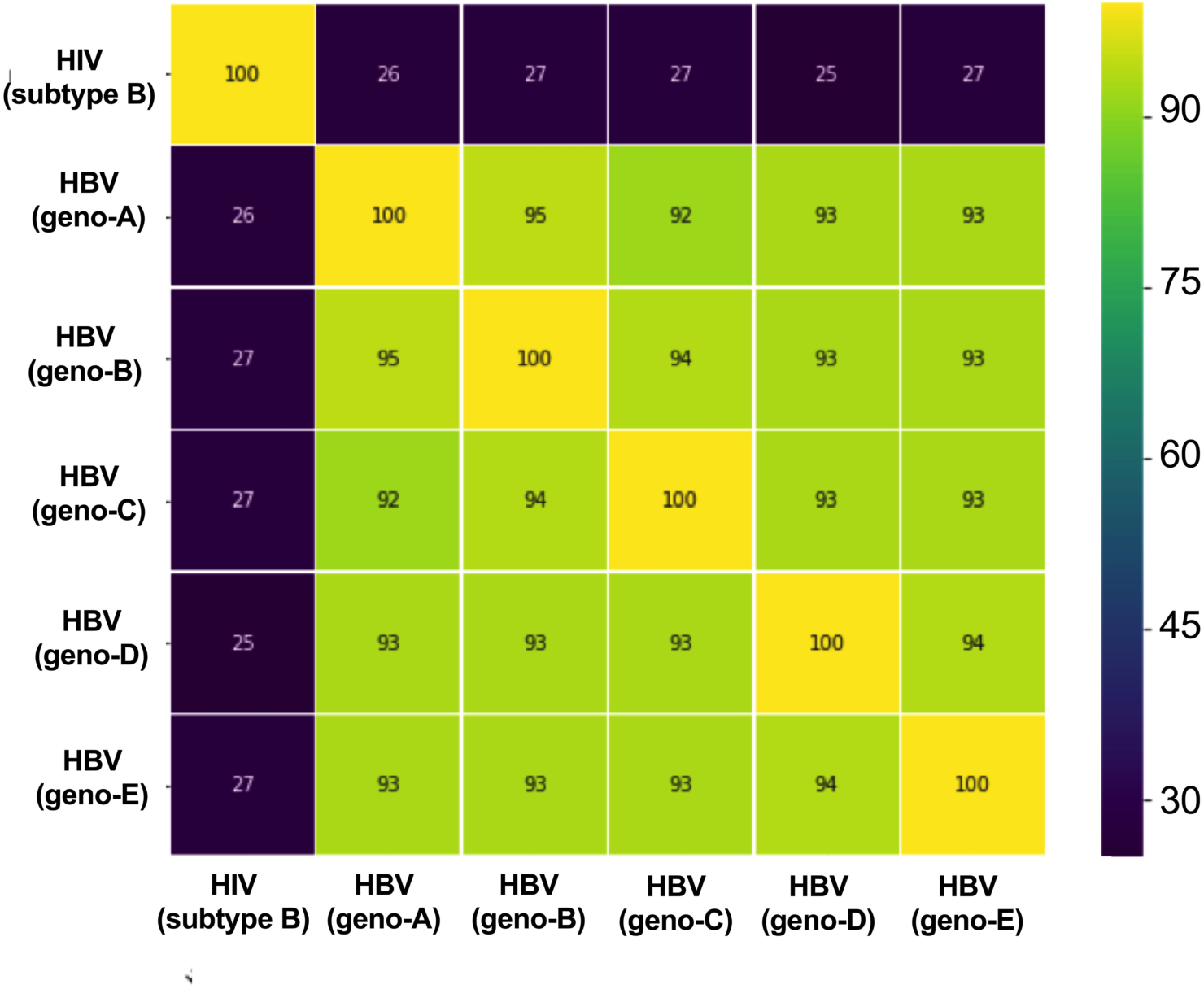
Heatmap showing identity comparison matrix of reference sequence alignment of HBV RT and HIV RT. Chart shows a comparison based on sequences downloaded from HIV sequence database (https://www.hiv.lanl.gov/) and Hepatitis B Virus Database (https://hbvdb.ibcp.fr/HBVdb/) and aligned using MAFFT version 7. HIV reference sequence is HIV HXB2 (K03455). HBV reference sequences are Geno A – FJ692557, Geno B -GU815637, Geno C – GQ377617, Geno D -KC875277, Geno E -GQ161817.

### Appraisal of putative sites of drug resistance

Recognising that there is sparse and varied evidence to support TDF RAMs, we divided our data into two categories. First, we generated a ‘long list’ of all polymorphisms that have been reported in association with TFV resistance, to summarise all the available data in the most inclusive way. We then refined this into a ‘short list’ including just those sites supported by the most robust evidence, based on reporting in ≥2 studies, and a combination of *in vivo* and *in vitro* evidence.

### Sequence analysis

To assess similarity between HIV and HBV RT, we downloaded HIV and HBV reference sequences from a publicly available repositories: HIV sequence database (15) and Hepatitis B Virus Database (16). We aligned amino acid RT sequences using MAFFT version 7 (17). The alignment illustrates regions of similarity and differences between HIV and HBV RT. We obtained RAMs associated with HIV resistance to TFV from Stanford University HIV drug resistance database (18).

### Structural analysis

The crystal structure for HBV RT has not been solved, However, the enzyme is homologous to HIV RT. In order to further assess the relevance of sites included in a ‘long list’ of all polymorphisms, we therefore considered evidence for a mechanistic influence by mapping HBV RAMs onto a previously solved crystal structure of HIV RT (PDB code 3dlk) (19) using ICM [http://www.molsoft.com/icm_pro.html].

## RESULTS

### Nature and quality of the evidence identified

We identified 15 studies that met our search criteria. Although some studies used hybrid methods, we classified them broadly as seven studies arising from clinical case reports (7,8,20–24), (one of these also presented in vitro evidence for drug resistance (8)), four from *in vitro* studies (25–27) and four from longitudinal studies of CHB (with or without HIV coinfection) (28–31). Studies were from Asia (7,8,20,22,27), Australia (31), Europe (21,24,26,28–30) and USA (23,25,32). Despite the high prevalence of HBV infection in Africa, and the widespread use of TFV for HIV across this continent, it is striking that no African data have been published to date. Eight studies reported HBV genotypes, representing genotypes A-G (8,20,23–25,28–30,32). Metadata for individual studies are provided in **Suppl Table 1**.

A detailed quality assessment of each individual reference is included in **Suppl Table 2**. Among the case reports, three were of high quality as they clearly described patients’ characteristics, clinical details, diagnosis, treatment and follow-up, and concluded with take away lessons (20–22). Three further case reports did not describe diagnostic or assessment methods (7,23,24), and one study did not describe post-intervention follow-up (8). The overall quality rating for four cohort studies was strong because participants selected represented the target population, characteristics of participants were clearly described, there was a clear hypothesis for the study, and inclusion/exclusion criteria were specified (28–31). Two studies had a weak rating because there was no description of the characteristics of participants and it was not clear to what extent participants were representative of the target population (25,32). Two natural experimental studies were not rated because the quality assessment questions were not applicable (26,27).

### Approach to defining resistance

Resistance was studied based on exposure to TDF in 13 studies and TAF in two studies; we therefore refer to TFV throughout the results section. TFV resistance was determined using a range of strategies, which can be summarised as follows:

i. A sequencing approach to identifying possible RAMs in HBV sequence isolated from individuals in whom viraemia was not suppressed by TFV therapy, undertaken in seven studies (7,20,22–24,28,31);
ii. *In vitro* assays to measure the effect of TFV on viral replication in cell lines, reported by three studies (25,26,32);
iii. Approaches (i) and (ii) in combination, applied in four studies (8,21,29,30);
iv. Approach (ii) combined with an animal model, described by one study (27).

In 12/15 studies, HBV mutations were reported in association with TFV resistance or reduced TFV susceptibility (8,20,21,24–30,32). In the remaining three studies, persistent viraemia was reported among individuals with chronic HBV infection while on TFV but either no RAMs were identified (23,31) or viral sequencing was not performed due to low viral load (22).

### Location and number of RAMs associated with TFV resistance

We generated a ‘long-list’ of 37 different sites of polymorphism, arising both within (n=15) and outside (n=22) enzymatically active sites in RT (Fig 1; **Suppl Table 3**). HBV mutations outside active sites of the enzyme occurred in combination with RAMs located within active sites, with the exception of A194T. Only two studies reported TFV resistance arising from the selection of a single mutation, S78T and A194T (21,26). S78T was defined by sequencing HBV from two individuals in whom viraemia was not suppressed by TDF, combined with *in vitro* assays (21), while A194T was only defined *in vitro* (26). In all other studies, ≥2 RAMs were required to confer TFV resistance (2 RAMs in four studies (25,27,28,32), 3 RAMs in one study (30), 5 RAMs in one study (8), and ≥8 RAMs in a further four studies (7,20,24,29)). This pattern supports the high genetic barrier to selection of TFV resistance.

We narrowed the list of 37 sites down to compile a ‘short-list’ of TFV RAMs that have been identified in ≥2 studies, regarding these sites as having the strongest evidence base (9 sites; **Table 1)**. The most frequently described RAMs were L180M (20,29,30), A181T/V (25,28,29,32), M204I/V (8,20,29,30), and N236T (25,28,32), which were all identified through sequencing and tested in *in vitro* assay to measure the effect of TDF on viral replication in cell lines. Among these, the M204 mutation (within the ‘YMDD’ motif) is well established in association with 3TC resistance, commonly arising in combination with substitutions at positions V173, L180 and A181, while N236 substitutions appear to be more specifically associated with reduced susceptibility to ADV and TFV (6). Mutations at sites 177, 194 and 249 may also be more specific to TFV resistance, having been less clearly reported in association with resistance to other agents (3,33). Polymorphisms at positions 80, 173 and 184 have been described as compensatory changes to allow the virus to accommodate the primary drug escape substitution (6). These mutations on their own may not be sufficient to mediate TVF resistance but may be a necessary compensatory contribution to combinations of mutations that underpin resistance.

Some reported polymorphisms associated with drug resistance represent wild type sequence in some genotypes (Y9H, F122L, H126Y, R153W/Q, F221Y, S223A, C256S, D263E, V278IV and A317S), and our assimilation shows more resistance in genotype D (**Suppl Table 4**). Most of these polymorphisms are located outside the active site of the enzyme, with the exception of position 256. The barrier to selection of TFV resistance may therefore be lower in certain genotypes, (Fig 1, **Suppl Table 4**). The same phenomenon has been described for HCV resistance, in which certain sub genotypes are predicted to be intrinsically resistant to certain direct acting antiviral agents due to the presence of resistance associated polymorphisms in the wild type sequence (34,35).

One study assessed the replication competence and susceptibility to TFV of mutated HBV clones *in vitro* and *in vivo* using mice models. The introduction of P177G and F249A mutations (substitutions in active sites of the RT enzyme) in HBV clones, resulted in a reduction in their susceptibility to TFV (27).

### RAMs occurring as minor quasispecies

There is limited evidence for the significance of TFV RAMs occurring as minor quasispecies. One study that performed ultra-deep pyrosequencing enrolled HIV/HBV co-infected individuals on (or about to start) TFV-containing ART, reporting minor variants present at <20% with mutations (V173, L180M, A181T/V and M204V) in 2/50 TFV naïve samples and 1/14 sample obtained from a TDF-experienced individual (31). One other study performed deep sequencing using Illumina on HBV clones (8), revealing that RAMs S106C, H126Y, D134E, M204I/V & L269I were predominant. Only one study reported sequencing the whole HBV genome, but this was undertaken following i*n vitro* introduction of RAMs into a clinically isolated virus (32), so does not provide any evidence of the association between TFV RAMs and other polymorphisms that might arise on the same viral haplotype.

### Duration of therapy and treatment compliance prior to detection of tenofovir resistance

Five studies reported the duration in which individuals were on TFV prior to treatment failure, with virological breakthrough occurring between 48 weeks and 48 months of therapy (48 weeks (20), 18 months (22), 20 months (29), 26 months (8), and 48 months (7,21)). Compliance was assessed in six studies, among which virological breakthrough despite good treatment compliance was reported in five (8,20,22,29,30), and one reported concerns with compliance (24). Quantification of drug levels in plasma supported good compliance in two studies (8,29).

### Comparison between HIV and HBV RT

HBV RT has been classified into subdomains which are further divided into regions A – G (36), which form the main catalytic core of the enzyme (Fig 1). Alignment of sequences of HBV and HIV RT demonstrates 25-27% homology between HBV and the HXB2 HIV reference sequence (Fig 2). Comparing sites that have been reported in association with drug resistance in HBV vs HIV (Fig 3, **Suppl Table 5**), we found that among our long-list of 37 HBV RAMs, two sites had identical substitutions in HIV RT (M204 and L229 in HBV (9,11), corresponding to M184 and L210 in HIV, respectively [https://hivdb.stanford.edu/dr-summary/resistance-notes/NRTI/], Fig 3). Other sites reporting an association with TVF resistance in HBV have substitutions that overlap with HIV RAMs, but not all of these are associated with TFV resistance, **Suppl Table 5**.

**Fig 3:**
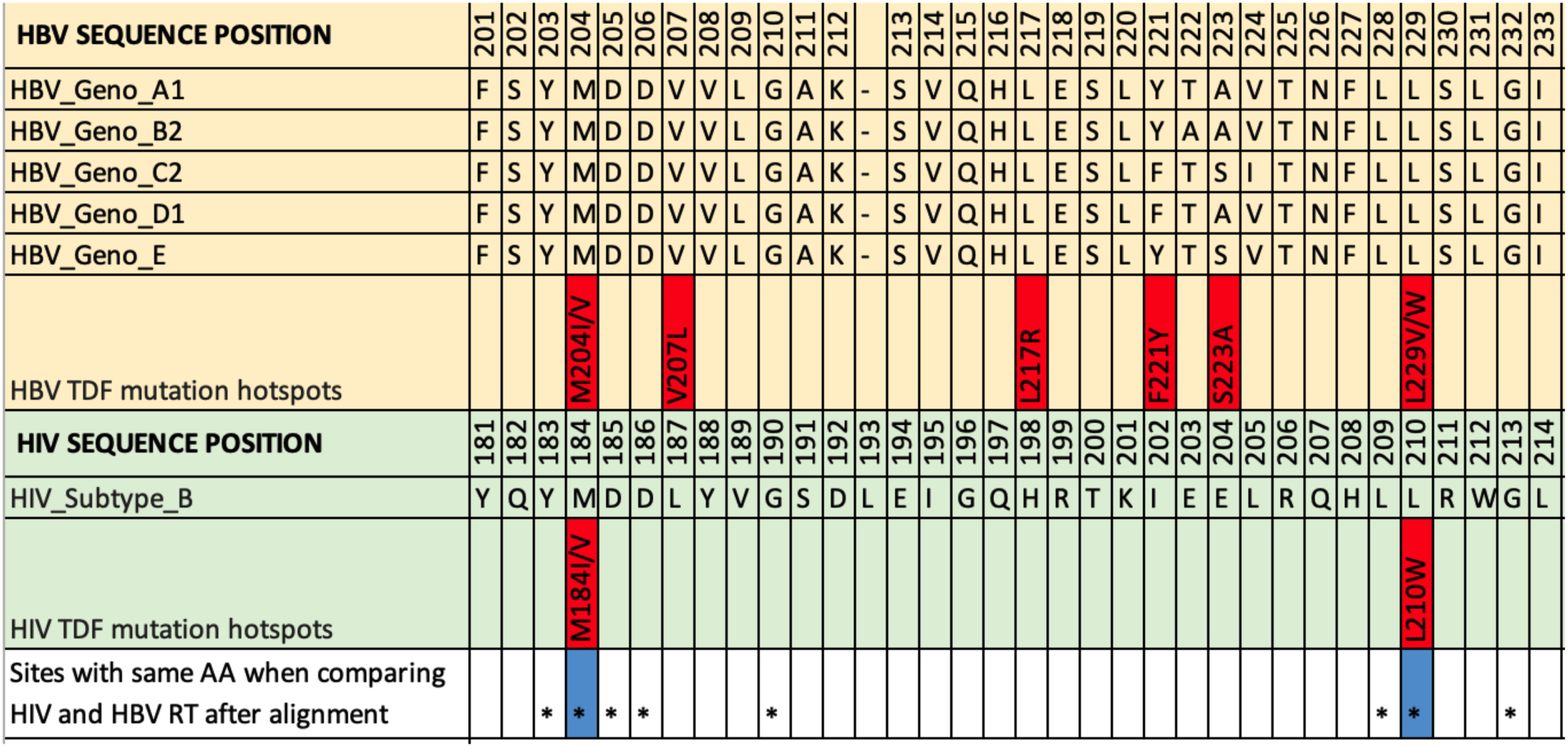
A section of the reference sequence alignment of HBV RT and HIV RT. Sequences downloaded from HIV sequence database at https://www.hiv.lanl.gov/ and Hepatitis B Virus Database at https://hbvdb.ibcp.fr/HBVdb/. Sequences were aligned using MAFFT version 7. HIV subtype B reference sequence is shown in light green (accession number K03455). HBV reference sequences are shown in yellow (Geno-A: FJ692557; Geno-B: GU815637; Geno-C: GQ377617; Geno-D: KC875277; Geno-E: GQ161817). Sites of TFV resistance are highlighted in red, based on the data assimilatad in this study. HIV tenofovir RAMs were obtained from the online Stanford Database https://hivdb.stanford.edu/dr-summary/resistance-notes/NRTI/. Sites marked * have the same amino acid in HIV and HBV RT after alignment, and those coloured blue also share TFV resistance mutations. This section is shown as it contains the only two homologous TFV RAMs that we have identified using this approach. Sequence alignments and RAMs throughout the whole RT protein is shown in Suppl Table 5. Note that in most cases, individual mutations are unlikely to be sufficient to mediate resistance, and a resistant phenotype arises only as a result of combinations of ≥2 polymorphisms.

Six established TFV RAMs in HIV RT (M41L, K65R, K70E, Y115F, Q151M and T215F/Y) (18) do not correspond to an equivalent mutation in HBV RT, although three of these HIV RAMs have an HBV RAM within 3 amino acids up-or down-stream in the equivalent sequence, suggesting there may be homology in the mechanism through which drug resistance is mediated.

We mapped HBV RAMs onto the crystal structure of the likely structurally-related HIV RT (pdb code 3dlk) in order to visualise their approximate 3D locations and infer possible functional consequences (Fig 4) (**Suppl Table 6**). The RAMs are primarily located within the ‘fingers’, ‘palm’, ‘thumb’ and ‘connection’ subdomains of the p66 polymerase domain of HIV RT, with the majority within the ‘palm’. A number of RAMs (e.g. V207, M204, F249) are spatially adjacent to the catalytically critical (and highly conserved) residues D110, D185 and D186 in HIV RT (D83, D204 and D205 in HBV), suggesting that these RAMs are highly likely to affected catalytic competency. Twenty-two HBV RAMs map to HIV RT residue positions which, if mutated, are likely to cause polymerase structure destabilisation, suggesting that many of these RAMs are likely to impact upon resistance.

**Fig 4:**
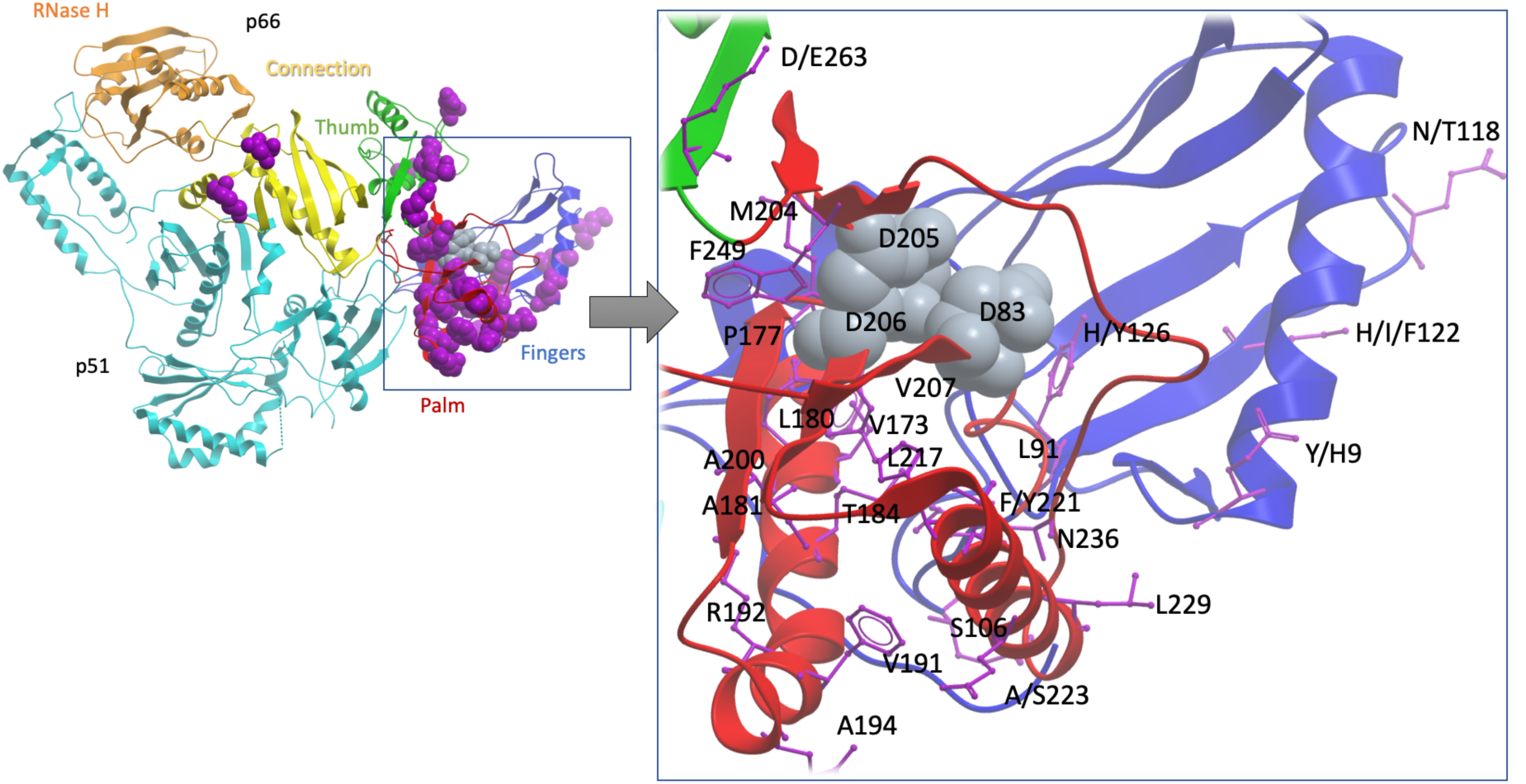
Cartoon to show the sites of TFV drug resistance polymorphisms, using the homologous crystal structure of HIV RT as a model. The sequence alignment of HBV was extended with HIV RT’s p66 domain and then projected onto a high-resolution HIV RT structure (pdb code 3dlk). Sub-domains of the HIV RT are coloured and annotated. Positions associated with resistance are scattered primarily throughout the finger and palm subdomains of the p66 domain (purple space-filled representations, left whole-molecule view, purple stick representation on the zoomed in view on the right). Three aspartate residues, D83, D205 and D206 (indicated by grey space-filled representation) form the catalytic triad of the enzyme and are shown as a point of reference. Of the 37 sites identified as potential TFV RAMs, 24 residues which are visible in the structure are labelled (using HBV numbering). This excludes seven putative HBV mutations at sites which do not have a homologous site in the HIV structure (sites 78, 80, 130, 134, 153, 163 and 256), and six sites which are beyond the end of the sequence of the solved crystal HIV structure (267, 269, 278, 317, 333 and 337). Figure produced using the ICM platform (http://www.molsoft.com/icm_browser.html). Note that in most cases, individual mutations are unlikely to be sufficient to mediate resistance, and a resistant phenotype arises only as a result of combinations of ≥2 polymorphisms.

## DISCUSSION

### Summary of key findings

TFV is a safe and effective treatment choice for CHB in the majority of cases, and large case series have not raised significant concerns about clinically significant drug resistance. However, it is important to consider the potential for the emergence of resistance, demonstrated by persistent viraemia on therapy and/or reduced virologic suppression *in vitro*. Based on existing evidence, TFV resistance seems likely to depend on the selection of suites of mutations (most commonly including L180M, A181V/T, M204I/V and/or N236T), overlapping with RAMs that allow escape from other NA drugs. There is also a suggestion that, rarely, single mutations can confer TFV resistance, best demonstrated for S78T.

Notably, the literature to date is limited and heterogenous, and there remains a lack of evidence about the frequency and likely impact of proposed TFV RAMs either within individual patients or at population level. At present, we have tackled this uncertainty by dividing our catalogue of polymorphisms into a ‘long-list’ (all reported RAMs) and a ‘short-list’ (RAMs with the best evidence-base of support).

Tools that have been designed to identify drug resistance may bias against detection of relevant mutations if they do not scrutinise all relevant sites that contribute to reducing TFV susceptibility. For example, ‘TRUGENE’ captures common HBV RAMs but does not include positions 78, 177, or 249 which may be pertinent to TFV resistance (37) and ‘geno2pheno hbv’ only lists one TFV mutation at position 236.

### Overlap of TFV RAMs with RAMs to other NA agents

RAMs L180M, M204I/V and A181T/V have been associated with resistance to 3TC, LdT and ETV (3,38–41); their reported association with TFV resistance is of concern in suggesting that prior NA exposure can increase the likelihood of cross-resistance to TFV. A study of HIV/HBV co-infected individuals demonstrated a decreased likelihood of HBV DNA suppression with TDF among individuals exposed to prolonged 3TC treatment, possibly due to accumulation of such mutations (42). A large study in China reported A181 and/or N236 substitutions in 11% of the population (40), which may underpin reduced susceptibility to TFV. The structural similarities between ADV and TFV, and similar interaction with HBV polymerase (1,2) explain why the ADV RAMs A181T/V and N236T are also reported to confer resistance to TFV (1,43).

Although TFV has been considered effective in the context of resistance to other NAs (44), the current evidence suggests that there may be common pathways to resistance (45). There is some evidence showing co-location of RAMs conferring resistance to different antiviral agents on the same viral haplotype (46). These findings suggesting cross-resistance are of concern, especially for settings in which there has been widespread use of NA therapy as a component of ART for HIV (3).

### Sites of TFV RAMs in HBV RT

Resistance to TFV can be explained by RAMs both within and outside the active site of the RT enzyme, some of which may have similar mechanisms to those described in HIV (9,36). The mechanism of resistance in most of these polymorphisms remains unknown, but may interfere with drug access to sites of activity through steric hindrance. Mutations within active sites of the enzyme may be associated with a higher fitness cost to the virus than mutations at other locations in the RT sequence, as they are more likely to interfere with the RT function. Some polymorphisms listed as RAMs may in fact represent compensatory mutations, that are co-selected in the presence of primary RAMs. For example, substitution at position 269 has been previously described as a compensatory mutation that restores impairments to RT function (47).

Currently, HBV genotyping is not routinely undertaken in clinical practice, so it is difficult to amass data for any potential relationship between resistance and viral genotype. However, there are some clues that genotype may be relevant. For example, C256S has been linked to TFV resistance, but S256 is wild type in genotype C, **(Suppl Table 4)**, suggesting that the genetic barrier to TFV resistance in genotype C might be lower than in other genotypes.

However, a study of >1000 individuals in China found no differences in drug resistance rates between Geno-B vs Geno-C infection (40). The identification of Y9H as a TFV RAM should be viewed with caution as H9 is frequently the wildtype residue, irrespective of genotype.

### Other factors associated with persistent vireamia

In addition to RAMs, there are other explanations for incomplete suppression of HBV viraemia on therapy (23,31), including a higher baseline HBV DNA level, positive baseline HBeAg status, history of 3TC exposure, a lower nadir CD4+ T cell count in the context of HIV coinfection, and high serum HBV RNA levels (42,48,49). Given that HBV DNA is inhibited in a dose-dependent manner (2), it is also possible that insufficient drug delivery to the infected hepatocyte could be the cause of persistent viraemia even in the absence of specific RAMs.

Incomplete adherence to therapy can also contribute to virological breakthrough (50). Two studies included in our review assessed treatment compliance by measuring drug concentration in plasma (8,29). Assessment of adherence in chronic HBV has been through the use of questionnaires (51), but these are subject to self-reporting bias. Evidence of potential TFV resistance may emerge when individuals with HIV/HBV coinfection are treated with a TFV-containing regimen leading to suppression of HIV but with sustained HBV vireamia (52).

It has been reported to take three years for 90% of HBV infected individuals to reach viraemic suppression on therapy (53), in contrast to HIV, in which 88% of patients suppress the virus within the first year of TDF-based treatment (54). In the studies we have reported in this review, persistent HBV viraemia on therapy could be due to the prolonged timeline for viraemic suppression; however, in most studies there was a reduction in viral load when TDF was initiated, with subsequent virological breakthrough that is more in keeping with the selection of resistance.

### Implications for patient management

There are not currently sufficient data about TFV RAMs to underpin robust universal guidelines for clinical practice. However, the evidence that we have gathered here can underpin some practical recommendations:

i. There is an urgent need for more HBV sequencing data, together with contemporaneous viral load measurements and clinical metadata to advance understanding of the relationship between viral sequence and treatment outcomes. Sequence repositories, databases and tools for sequence analysis should regularly review the evidence for TDF RAMs in order to highlight all sites that may be significant in mediating resistance.
ii. In the context of failure of viraemic suppression in a patient prescribed therapy, assessing and supporting drug compliance is crucial, ideally together with viral sequencing.
iii. Therapeutic failure of TFV – whether in the presence or absence of known RAMs – should lead to an expert clinical decision about switching therapy or combining agents. The presence of recognised RAMs, especially when in combination, may support a change of therapy, considering ETV or combination therapy, although guidance and options are currently limited.
iv. If there is an ongoing emergence of data to suggest TFV resistance, there will be a need for expert guidelines to include practical recommendations in order to unify clinical approaches. HIV guidelines should take an active stance on incorporating recommendations for those with HBV co-infection, particularly if dual therapy regimens are adopted.
v. Evidence of TFV resistance highlights the need for development of robust novel direct acting antivirals and immune therapies for HBV.

### Caveats and limitations

There is sparse literature on HBV resistance to TFV, and studies are of varying quality. While there is a high genetic barrier to selection of TFV resistance, it is likely that there is under-reporting of cases of resistance, particularly in low/middle income settings in which routine monitoring of HBV VL on treatment is not undertaken. It can be difficult to infer the impact of common polymorphisms on drug resistance phenotype; for example, it is plausible that M204I/V may be enriched among TFV resistant strains simply as a ‘footprint’ of prior exposure to 3TC.

Most studies to date have used Sanger sequencing, and it is possible that significant minority variants may be under-represented, as suggested by one report in which phenotypic TFV resistance was associated with RAMs in <20% of minor variants (31). Low HBV DNA viral loads are a further barrier to sequencing, and bias existing data towards samples from individuals with high viral loads, in which the full spectrum of relevant RAMs may not occur. It is therefore important to invest in deep sequencing platforms that offer the opportunity to explore the full landscape of HBV variants isolated from an infected individual, and to improve sensitivity of sequencing methods including both Sanger and ‘next-generation’ approaches. Some sequencing methods, such as Oxford Nanopore Technologies, can generate long reads that allow reconstruction of complete viral haplotypes, providing improved certainty about linkages between sites (55). To be able to undertake an appropriate haplotypes analysis would require datasets with robustly phenotyped patients (displaying clinical evidence of drug resistance) together with full length viral sequence data; such datasets have not been generated to date but are an important long-term aim.

We recognise the limitations of drawing direct comparisons between HIV and HBV RT, given the limited (<30%) sequence homology between the two enzymes, and the finding that only 2/37 sites associated with TVF resistance in HBV are homologous RAMs in HIV. This highlights a need for future work to solve the crystal structure of HBV RT.

## Conclusions

We have assimilatad emerging evidence for HBV polymorphisms that reduce susceptibility to TFV, also acknowledging the potential influences of other viral and host factors in cases of persistent viraemia on therapy. While the genetic barrier to resistance is high, evidenced by the large number of mutations that typically have to be selected to produce resistance, of concern is the overlap with other NA resistance mutations, and the instances in which individual amino acid polymorphisms may be sufficient to produce phenotypic resistance. Enhanced studies representing larger numbers of patients, tracking longitudinal viral sequence changes, and monitoring viral suppression over time are needed. In addition, the evolution of better *in vitro* models will support experiments to investigate the effect of individual and combined RAMs. In order to optimise the use of NA therapy as a tool in driving advancements towards elimination at a population level, improved insights into drug resistance are essential. If resistance emerges as a substantial clinical problem, there will be a need for consideration of synergistic drug regimens, new agents that inhibit a target other than viral RT, and for the development of new therapeutic strategies that can bring about cure.

## Data Availability

Data availability
All data generated or analysed during this study are included in this published article (and its Supplementary Information files).

https://figshare.com/s/7dcfa78cc5ed14094d6f

## AUTHORS CONTRIBUTIONS

JM and PCM conceived the study. JM performed literature review. JM, PAB, BDM, PCM analysed the data. JM and PCM wrote the manuscript. All authors revised and approved the final manuscript.

## TRANSPARENCY DECLARATION

Nothing to declare.

## SUPPLEMENTARY DATA

Supporting data files can be accessed on-line using the following link: https://figshare.com/s/7dcfa78cc5ed14094d6f

On acceptance of the manuscript for publication, the following permanent DOI will be made available: 10.6084/m9.figshare.8427746.

**Suppl Fig 1: Pharmacokinetics of tenofovir disoproxil fumarate (TDF), tenofovir alafenamide (TAF) and adefovir dipivoxil (ADV)**. Orange rectangle represents the gut, showing amount of drug absorbed in mg; yellow rectangle represents plasma, in which TDF and ADV are converted to the active molecules, tenofovir and adefovir respectively. Green rectangle represents hepatocytes where the active drug is incorporated into growing HBV DNA to stop viral replication.

**Suppl Fig 2:** PRISMA flow diagram to illustrate the identification of studies for inclusion in a systematic review of tenofovir resistance in HBV infection (provided in pdf format).

**Suppl Table 1: Metadata for 15 studies reporting TDF resistance in HBV infection** identified from a systematic literature review.

**Suppl Table 2: Quality assessment for 15 studies reporting TDF resistance in HBV infection**. Studies identified from a systematic literature review.

**Suppl Table 3: List of 37 tenofovir RAMs in HBV Reverse Transcriptase (RT) reported in 15 studies identified through a systematic literature review** (provided in xls format).

**Suppl Table 4: List of TFV RAMs in HBV, showing consensus residue at each position according to HBV genotype, and genotype-specific resistance where this has been reported**. List of RAMs obtained from 12 studies included in the review. HBV reference sequences used are the same as for Fig 1. Grey colour highlights the sites that differ from majority consensus. Ticks represent RAMs situated in sites where there is a difference (at consensus level) between genotype sequences. RAMs are listed with the wild-type amino acid first, followed by the numbered position in RT sequence, and the amino acid substitution that is associated with resistance. TFV = tenofovir, RAMs = resistance associated mutations, HBV = hepatitis B virus.

**Suppl Table 5: Reference sequence alignment of HBV RT and HIV RT**. Sequences downloaded from HIV sequence database online at https://www.hiv.lanl.gov/ and Hepatitis B Virus Database online at https://hbvdb.ibcp.fr/HBVdb/. Alignment undertaken using MAFFT version 7. HIV subtype B reference sequence is shown in light green (accession number K03455). HBV reference sequences (Geno-A: FJ692557; Geno-B: GU815637; Geno-C: GQ377617; Geno-D: KC875277; Geno-: GQ161817 shown in light yellow. TFV RAMs are highlighted in red. HIV TFV RAMs were obtained from Stanford Database (https://hivdb.stanford.edu/dr-summary/resistance-notes/NRTI/). HBV TFV RAMs were obtained from 12 studies included in the review. Sites with the same amino acid at homologous sites are indicated with asterisks (*); among these sites, those that are also shared TVF RAMs are shaded in light blue.

**Suppl Table 6: Mapping of RAMs to the HIV RT structure**. RAMs were mapped onto the high-resolution structure of HIV RT to infer their possible functional consequences in terms of structure.

## ABBREVIATIONS

3TC: lamivudine
ADV: Adefovir
ART: antiretroviral therapy
cccDNA: covalently closed circular DNA
CHB: chronic hepatitis B virus infection
ETV: entecavir (ETV)
HBsAg: Hepatitis B surface antigen
HBV: Hepatitis b virus
HIV: human immunodeficiency virus
LdT: telbivudine
NA: Nucleos(t)ide analogue
RAM: Resistance Associated Mutation
RT: reverse transcriptase
TAF: tenofovir alafenamide
TDF: tenofovir disoproxil fumarate
TFV: tenofovir (the active component of both TAF and TDF)
YMDD: tyrosine methionine aspartate aspartate motif in HBV RT

